# Trends, correlates, and recent pattern of antibiotic misuse in acute rotavirus diarrhea in urban and rural Bangladesh

**DOI:** 10.1101/2024.12.23.24319535

**Authors:** Kazi Nazmus Saqeeb, S. M. Tafsir Hasan, Soroar Hossain Khan, Md Alfazal Khan, ASG Faruque, Tahmeed Ahmed

## Abstract

**Background:** The indiscriminate use of antibiotics in pediatric populations has emerged as a critical global public health concern. A notable example of this is the misuse of antibiotics for treating rotavirus infections, particularly in developing countries. Despite this, there is a dearth of comprehensive research from this region. To address this gap, this study systematically examined the trends and factors associated with antibiotic misuse for acute rotavirus diarrhea among children aged 6-23 months in Bangladesh over a 15-year period. The study also explored sources of prescription, and types of antibiotics utilized in both urban and rural settings of Bangladesh.

**Methods:** Data from the icddr,b’s Diarrhea Disease Surveillance System (DDSS) were analyzed for 4870 children with laboratory-confirmed acute rotavirus diarrhea treated at Dhaka (urban) and Matlab (rural) hospitals between 2004 and 2018. Relevant sociodemographic and epidemiological data was obtained from the database. To assess changes in antibiotic use over the years chi-square test for trend was employed. Separate logistic regression models specific to each site were developed to identify factors linked to antibiotic use in cases of rotavirus diarrhea.

**Results:** Over the study period, the percentage of children with rotavirus diarrhea treated with antibiotics significantly rose in both urban (from 43% to 75.5%) and rural (from 35% to 69%) settings (p<0.001). In urban areas, a majority of children (57.5%) received antibiotics at a physician’s clinic for their illness, whereas almost all rural children (86.3%) obtained antibiotics from a pharmacy before being treated at icddr,b. Macrolides were identified as the most frequently prescribed antibiotics (46.6% in urban areas and 38% in rural areas). The urban regression model, revealed that factors such as severity of illness (OR = 1.8; 95% CI 1.5, 2.2), mother’s education (OR = 2.1; 95% CI 1.6, 2.8), father’s education (OR = 1.8; 95% CI 1.4, 2.3), household monthly income > $100 (OR = 1.5; 95% CI 1.2, 1.9), and the distance from home to the nearest health facility (OR = 1.4; 95% CI 1.1, 1.9) were all significantly positively correlated with the use of antibiotic among children suffering from rotavirus diarrhea. Similar results were observed in the rural regression model.

**Conclusions:** The increasing trend of antibiotic misuse for rotavirus diarrhea in Bangladesh, coupled with the tendency of healthcare providers to prescribe antibiotics inappropriately and the higher incidence of misuse among affluent, educated families, is alarming. Future research is therefore imperative to elucidate the hindrances and catalysts to the prudent administration of antibiotics across diverse societal groups, encompassing both healthcare personnel and family members.

## Introduction

Prescription and nonprescription use of antibiotics without proper clinical indication have appeared as a serious public health concern in recent times. In the USA, a high-income country with a seemingly developed health system, each year, about 47 million antibiotic prescriptions are dispensed inappropriately by physicians for viral illnesses (1). In low-and middle-income countries (LMICs), more than half of all antibiotics are sold in pharmacies without prescriptions from registered physicians/pharmacists (2). This growing practice of unnecessary antibiotic use in LMICs poses a substantial financial burden on the overall healthcare budget (3–5). In addition to the economic burden, individuals treated inappropriately with antibiotics are being exposed to the adverse health effects of these drugs. Antibiotics have been found to be the second most common cause of adverse drug reactions, including death (6). Moreover, it is also evident that developing countries, due to their widespread misuse of antibiotics, are contributing significantly to the worldwide rise of antibiotic-resistant pathogens (7). Globally, drug-resistant infections are causing 700,000 deaths per year, which is projected to increase to 10 million by 2050 (8).

Since viral infections are quite common during childhood, children often fall victim to the practice of antibiotic misuse, and systemic antibiotics are frequently prescribed. Studies from LMICs have shown that more than half of all children suffering from a viral infection received an antibiotic, and one-fifth without any prescription (9–12). Rotavirus diarrhea is one of the common viral infections of early childhood and has been identified as the primary cause of childhood diarrhea across all continents. It is estimated that worldwide, 95% of children under the age of 5 years experience a rotavirus infection regardless of socioeconomic status or geographic condition. By affecting an estimated 140 million children aged between six months to two years and causing 215,000 deaths per year, it becomes one of the most significant viral infections in early childhood (13,14). In rural Bangladesh, one-third of the total hospitalized cases of childhood diarrhea can be attributed to rotavirus infection (15). Most rotavirus infection usually takes place in the latter half of infancy after the initiation of complementary feeding (15–18). Since rotavirus diarrhea involves fluid loss, rehydration with oral rehydration solution (ORS) is the recommended treatment modality; the use of antibiotics is not advocated by the World Health Organization (WHO). In contrast to the recommendation, more than 40% of children with acute watery diarrhea receive unnecessary antibiotics in LMICs (19). MAL-ED, a large multicounty prospective cohort study, found that 57% of episodes of watery diarrhea in children below 2 years were treated with antibiotics in Dhaka, the capital of Bangladesh (20). Furthermore, Ahmed et al. showed that, in two different parts of Bangladesh, children with rotavirus infection had 1.5-1.6 times higher odds of receiving antibiotics compared to those suffering from other diarrheal illnesses (21,22).

There is a paucity of data from Bangladesh on long-term trends, patterns, determinants, and urban-rural differentials regarding the inappropriate use of antibiotics for rotavirus diarrhea. In this context, this study investigated the longitudinal trends in antibiotic misuse, associated factors, advocating sources, and types of antibiotics used among 6-23 months old children with acute rotavirus diarrhea in urban Dhaka and rural Matlab of Bangladesh.

## Methods

### Settings

The International Centre for Diarrhoeal Disease Research, Bangladesh (icddr,b) operates two specialized diarrheal disease hospitals in Dhaka (urban) and Matlab (rural). These facilities provide free treatment to approximately 300,000 patients annually, with over half being children under five, including a significant number with rotavirus diarrhea.

icddr,b established the Diarrheal Disease Surveillance System (DDSS) at both hospitals to collect data on patients. The urban DDSS in Dhaka randomly samples 2% of patients coming to Dhaka hospital, while the rural DDSS in Matlab enrolls all patients coming from the Matlab Health and Demographic Surveillance System (HDSS) area of icddr,b. Informed consent is obtained from patients or their guardians before enrollment. A structured questionnaire is administered to gather information on sociodemographic factors, disease-related epidemiologic factors, and medication use before coming to the hospital. Stool samples are collected and tested for pathogens, including rotavirus, using Enzyme-linked immunosorbent assays (ELISA).

The DDSS research assistant inquires about medication use, including antibiotics. Antibiotics are identified through drug inspection, prescription review, or recall. Common antibiotics are listed in the questionnaire, with an “others” category for unlisted drugs. The names of “others” category antibiotics were not routinely recorded until 2018, when they began to outnumber the listed antibiotics. The details of the population, data collection, and laboratory procedures of DDSS have been described elsewhere (23).

### Study population and data sources

Children aged 6-23 months presenting with acute rotavirus diarrhea to Dhaka and Matlab hospitals and enrolled in the DDSS between January 2004 and December 2018 constituted the study population. Data were retrieved from the DDSS database. The final sample included 2,864 urban and 2,006 rural children. As our aim was to investigate antibiotic misuse among children with rotavirus diarrhea without any comorbidities that might require an antibiotic, we excluded children who had diarrhea caused by rotavirus plus other pathogens, duration of the present episode of 14 days or longer, any previous episode of diarrhea in the past month, or history of any another illness in the preceding month. We also excluded those with missing information or received medications that could not be identified or confirmed (**Supp. Fig. 1 and 2**).

### Statistical analysis

We analyzed study population characteristics using proportions for categorical variables and medians with interquartile ranges for continuous variables. Demographic comparisons between groups were conducted using χ² or Fisher’s exact tests (categorical) and Student’s t-tests or Mann-Whitney U tests (continuous) as appropriate. The source of antibiotic advice, either from physicians or pharmacies, was compared between sites using χ² tests. We also examined the frequency and relative frequency of antibiotics used in 2018 at both sites. A chi-square test for trend assessed the linear trend of antibiotic use over 15 years.

Several sociodemographic and epidemiological factors were selected to identify associations with antibiotic use in acute rotavirus diarrhea based on available data and literature. These factors included sex, nutritional status (stunting and wasting), parental literacy, family income, distance to the nearest health facility, stool frequency in the past 24 hours, and the duration of diarrhea prior to admission. Acute diarrhea was defined as three or more loose, liquid, or watery stools in 24 hours, lasting less than 14 days (24,25). Stunting and wasting were defined based on length-for-age and weight-for-length Z scores using WHO standards (26).

Simple and multivariable logistic regression models were constructed for urban and rural sites to identify factors associated with antibiotic use in acute rotavirus diarrhea. Variables with a P-value less than 0.2 in bivariate models were included in multivariable models (27). Multicollinearity was assessed using the variance inflation factor. The strength of association was expressed as odds ratios (OR) with 95% confidence intervals. A P-value less than 0.05 was considered statistically significant. Data analysis was performed using IBM SPSS Statistics, Windows (Version 21; SPSS Inc., Chicago, IL, USA).

### Ethics statement

The Diarrheal Disease Surveillance System (DDSS) is an integral component of the ongoing operations at icddr,b’s Dhaka and Matlab hospitals. Informed verbal consent was obtained from all prospective patients or their legal guardians/caregivers at the time of enrollment. This consent was documented by checking the appropriate box on the questionnaire and was reviewed with the participants or their parents for confirmation. Participants were properly informed of the voluntary nature of their participation and their right to withdraw at any time. The confidentiality of the information collected was assured, and participants were informed that future analyses and publications would be conducted anonymously. The collected information was explained to be used for improving patient care. This surveillance process has been approved by the Research Review Committee (RRC) and Ethics Review Committee (ERC) of icddr,b (23).

## Results

Among 4,870 children who fulfilled our selection criteria, 2,864 (59.0%) were from the urban, and 2,006 (41.0%) were from the rural surveillance site. The majority of the children were boys (62.7% in urban and 62.5% in rural), had a literate father (84.6% in urban and 87.6% in rural), had a literate mother (87.8% in urban and 92.3% in rural), belonged to a family with monthly income more than 100 US$ (70.6% in urban and 60.6% in rural). The rate of wasting was similar between urban and rural children at 16.2% and 16.9%, respectively. The majority of the urban children were from households situated more than 5 miles away from the health facility (88.2%) and had a frequency of diarrhea >10 times in the last 24 hours (61.7%). On the contrary, only 33.3% of rural children living more than 5 miles away from the health facility, which is reflected in their disease status, and only 39.7% had a history of more than 10 episodes of diarrhea in 24 hours before arrival to the hospital. Of the urban children, 68.2% took antibiotics for the illness before coming to the hospital. In the rural sites, the rate of antibiotic use was 44.9% (**Table 1**).

**Table 1.** Characteristics of 6-23 months old children with rotaviral diarrhea, N = 4870.

| Characteristics | Urban (2864)<br>n (%) | Rural (2006)<br>n (%) | P value |
| --- | --- | --- | --- |
| Age, months; median (IQR) | 10 (8,13) | 11 (8,15) | <0.001 |
| Sex, male | 1797 (62.7) | 1254 (62.5) | 0.880 |
| Literate father | 2415 (84.3) | 1758 (87.6) | 0.001 |
| Literate mother | 2516 (87.8) | 1852 (92.3) | <0.001 |
| Distance of health facility from home, >5 miles | 2527 (88.2) | 668 (33.3) | <0.001 |
| Monthly income, >100US\$ | 2023 (70.6) | 1215 (60.6) | <0.001 |
| Stunting | 548 (19.1) | 440 (21.9) | 0.019 |
| Wasting | 465 (16.2) | 339 (16.9) | 0.556 |
| No. of stools in past 24 hours, >10 times | 1767 (61.7) | 797 (39.7) | <0.001 |
| Received antibiotic | 1954 (68.2) | 900 (44.9) | <0.001 |

In the urban site, the proportion of children who received antibiotics before coming to the hospital increased over the years from 43% in 2004 to 76% in 2018 (chi-square for trend p<0.001). In the rural site, the proportion of children who received antibiotics before coming to the hospital showed an even greater proportional upward trend, increasing from 35% in 2004 to 69% in 2018 (chi-square for trend p<0.001). However, there was a drop in injudicious antibiotic use in Matlab from 2008 to 2012. The rate of antibiotic use decreased in the year 2008 (28.9%) from 46.3% in the previous year, remained low until 2012 (22.7%), and then sharply rose again in 2013 (54.8%). This rising trend was maintained until the end of the study period in 2018 when it was at its highest (68.8%) (**Fig. 1**).

**Figure 1:**
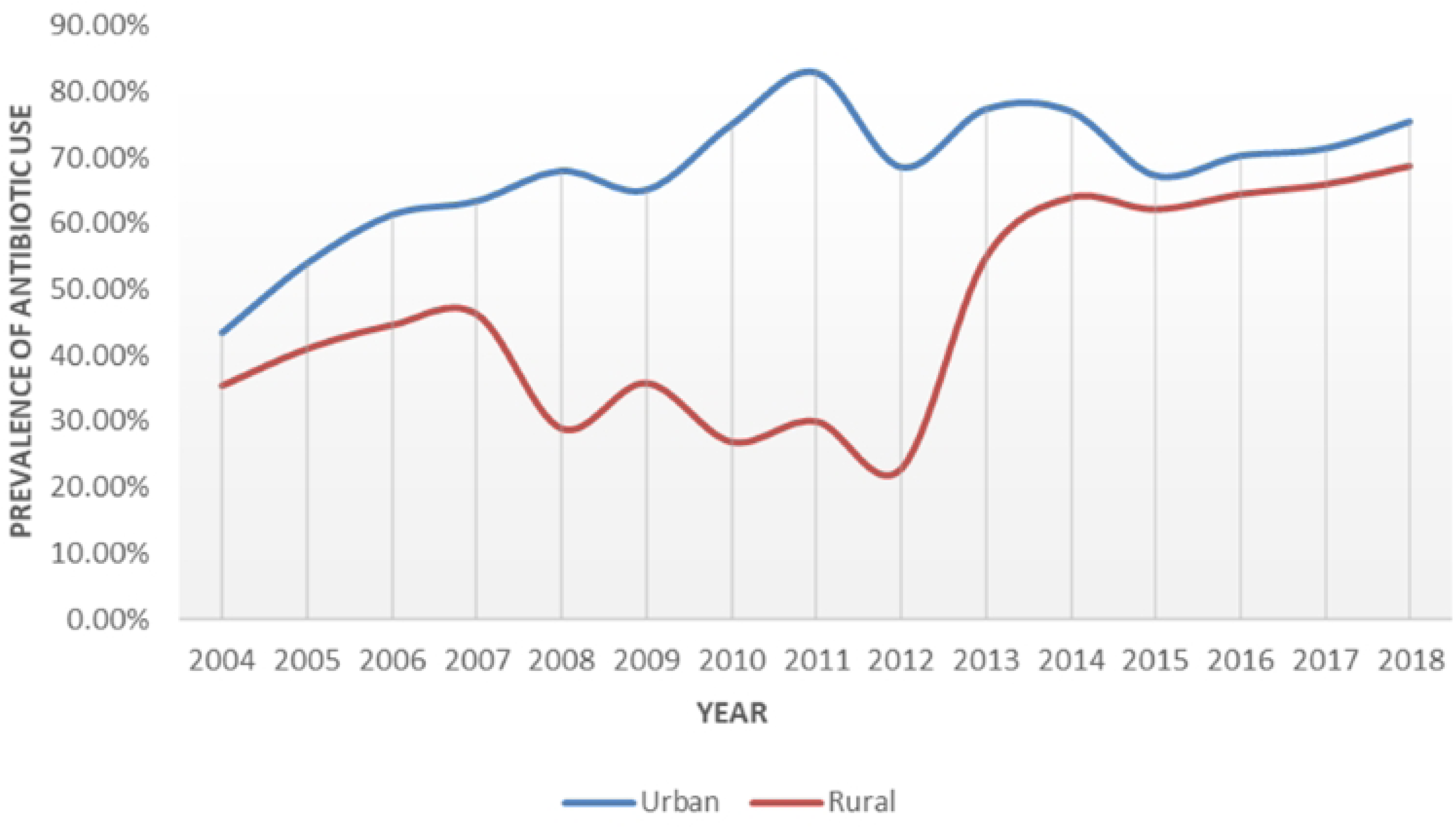
15 years trends of antibiotic use among 6-23 months old children with rotavirus diarrhea.

Bivariate analyses among urban children with acute rotavirus diarrhea found an association of antibiotic use with having a literate father or mother, stunting, wasting, monthly family income of more than 100 US$, living more than 5 miles away from the nearest health facility, passage of stool more than 10 times in the past 24 hours, duration of diarrhea prior to admission and admission time period. In the multivariable model, odds of receiving antibiotics in acute rotavirus diarrhea were higher in children who had a literate father (OR=1.8; 95% CI 1.4, 2.3), had a literate mother (OR=2.1; 95% CI 1.6, 2.8), had a family income of more than 100 US$ (OR=1.5; 95% CI 1.2, 1.9), lived more than 5 miles away from the nearest health facility (OR=1.4; 95% CI 1.1, 1.9), passed stool more than 10 times in the past 24 hours (OR=1.8; 95% CI 1.5, 2.2) and had diarrhea for longer duration (in hours) (OR=1.02; 95% CI 1.01, 1.02). Compared to 2004-2006, children admitted in the following years were more likely to receive antibiotics (**Table 2**).

**Table 2.**
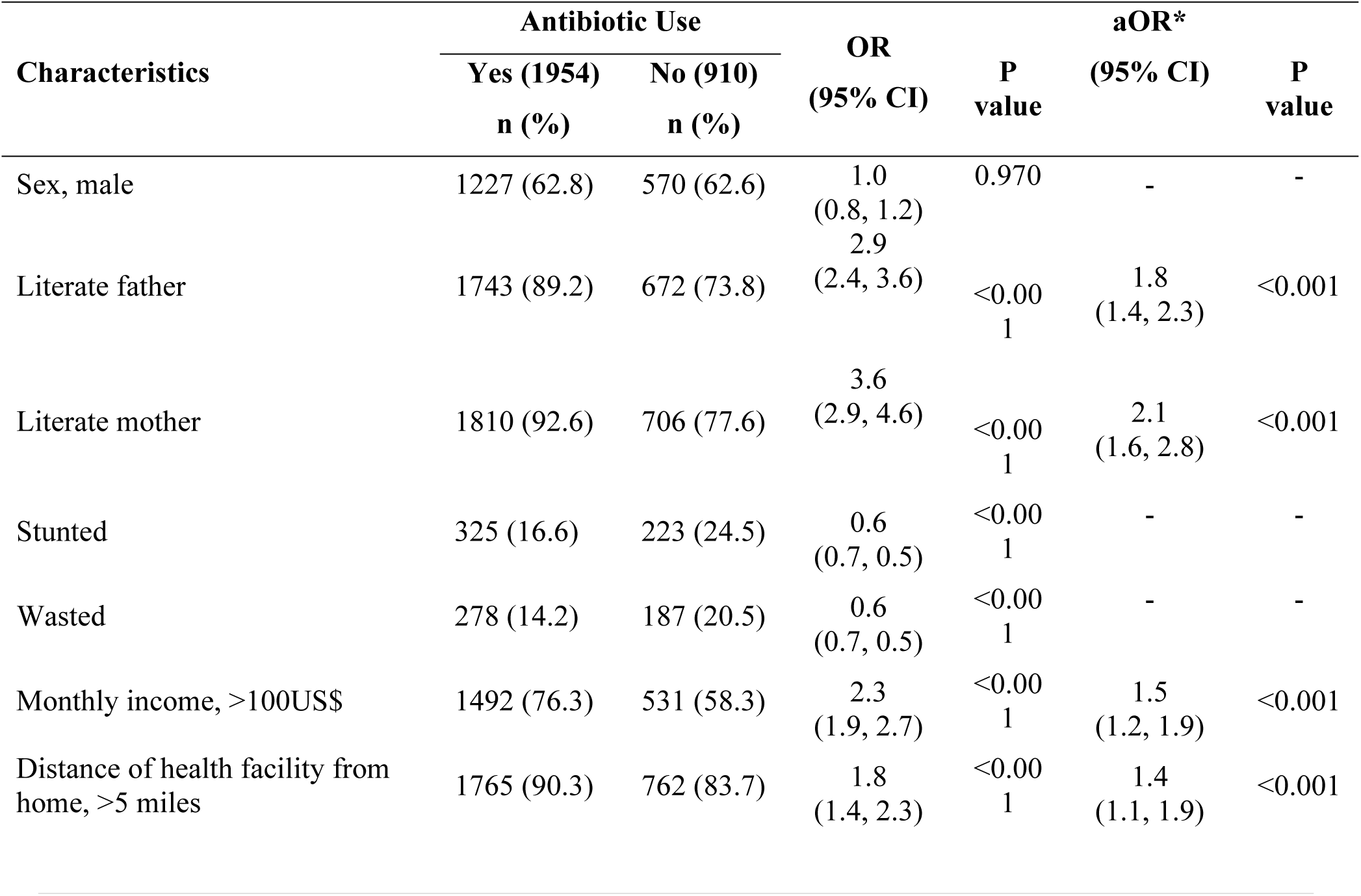
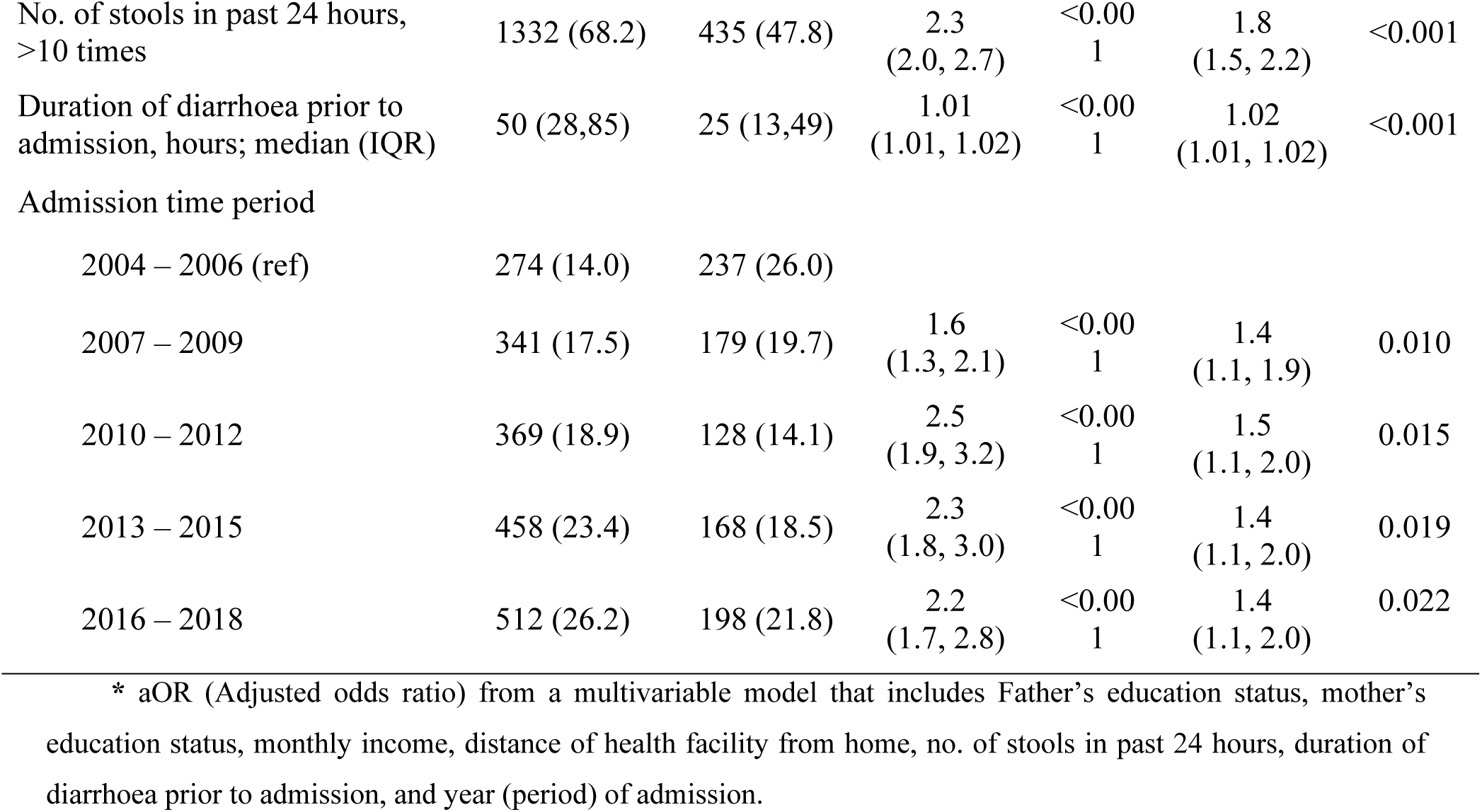
Correlates of antibiotic use among 6-23 months old urban children with rotavirus diarrhea, N= 2864.

Similarly, for the rural site, in bivariate analyses, literacy of father and mother, stunting, wasting, family income, living >5 miles away from the nearest health facility, passage of stool >10 times in the last 24 hours, duration of diarrhea prior to admission and admission time period were associated with antibiotic use in rotavirus diarrhea. In the multivariable model, odds of receiving antibiotics in acute rotavirus diarrhea were seen higher in children who had a literate father (OR=1.6; 95% CI 1.1, 2.3), had a literate mother (OR=1.6; 95% CI 1.1, 2.3), had a family income of more than 100 US$ (OR=1.7; 95% CI 1.3, 2.2), lived >5 miles away from the nearest health facility (OR=1.4; 95% CI 1.1, 1.7), passed stool >10 times in the last 24 hours (OR=1.8; 95% CI 1.4, 2.2) and had diarrhea for longer duration (in hours) (OR=1.02; 95% CI 1.01, 1.02). Children admitted in between 2007-2009 (OR=0.7; 95% CI 0.5, 0.9) and 2010-2012 (OR=0.5; 95% CI 0.3, 0.7) were less likely to have antibiotics (**Table 3**).

**Table 3.** Correlates of Antibiotic use among 6–23-month-old rural children with rotavirus diarrhea, N= 2006.

| Characteristics | Antibiotic Use |  | OR<br>(95% CI) | P value | aOR*<br>(95% CI) | P<br>value |
| --- | --- | --- | --- | --- | --- | --- |
|  | Yes (900)<br>n (%) | No (1106)<br>n (%) |  |  |  |  |
| Sex, male | 563 (63.0) | 691 (62.0) | 1.0<br>(0.8, 1.2) | 0.991 | - | - |
| Literate father | 832 (92.0) | 926 (84.0) | 2.4<br>(1.8, 3.2) | <0.001 | 1.6<br>(1.1, 2.3) | 0.015 |
| Literate mother | 863 (96.0) | 989 (89.0) | 2.8<br>(1.9, 4.0) | <0.001 | 1.6<br>(1.1, 2.3) | 0.050 |
| Stunted | 158 (18.0) | 282 (25.0) | 0.6<br>(0.7, 0.5) | <0.001 | - | - |
| Wasted | 118 (13.0) | 221 (20.0) | 0.6<br>(0.7, 0.5) | <0.001 | - | - |
| Monthly income, >100US\$ | 655 (73.0) | 560 (51.0) | 2.6<br>(2.1, 3.1) | <0.001 | 1.7<br>(1.3, 2.2) | <0.001 |
| Distance of health facility from<br>home, >5 miles | 345 (38.0) | 323 (29.0) | 1.5<br>(1.2, 1.8) | <0.001 | 1.4<br>(1.1, 1.7) | 0.002 |
| No. of stools in past 24 hours, >10<br>times | 471 (52.0) | 326 (29.0) | 2.6<br>(2.2, 3.2) | <0.001 | 1.8<br>(1.4, 2.2) | <0.001 |
| Duration of diarrhoea prior to<br>admission, hours; median, (IQR) | 38 (23,64) | 20 (12,34) | 1.02<br>(1.01, 1.03) | <0.001 | 1.02<br>(1.01, 1.02) | <0.001 |
| Admission time period |  |  |  |  |  |  |
| 2004 – 2006 (ref) | 151 (16.8) | 229 (20.7) |  |  |  |  |
| 2007 – 2009 | 209 (23.2) | 375 (33.9) | 0.8<br>(0.6, 1.1) | 0.215 | 0.7<br>(0.5, 0.9) | 0.020 |
| 2010 – 2012 | 91 (10.1) | 250 (22.6) | 0.5<br>(0.4, 0.7) | <0.001 | 0.5<br>(0.3, 0.7) | <0.001 |
| 2013 – 2015 | 186 (20.7) | 119 (10.8) | 2.4<br>(1.7, 3.2) | <0.001 | 1.2<br>(0.8, 1.7) | 0.360 |
| 2016 – 2018 | 263 (29.2) | 133 (12.0) | 2.9<br>(2.2, 4.0) | <0.001 | 1.3<br>(0.9, 1.9) | 0.122 |

\* aOR (Adjusted odds ratio) from a multivariable model that includes Father's education status, mother's education status, monthly income, distance of health facility from home, no. of stools in past 24 hours, duration of diarrhoea prior to admission, and year (period) of admission.

In 2018, macrolides were the predominant class of antibiotic used both in urban (45.9%) and rural (39.7%) areas, followed by Quinolones (36.1%) in the urban area and Nitroimidazole (27.0%) in the rural area (**Table 4**).

**Table 4.** Pattern of antibiotic use in 6-23 months old children with rotavirus diarrhea in 2018.

| Antibiotic | Urban (133)<br>(n%) | Rural (63)<br>(n%) |
| --- | --- | --- |
| **Macrolide** |  |  |
| Azithromycin | 52 (39.1) | 18 (28.6) |
| Erythromycin | 9 (6.8) | 7 (11.1) |
| **Quinolone** |  |  |
| Ciprofloxacin | 45 (33.8) | 15 (23.8) |
| Levofloxacin | 1 (0.8) | - |
| Nalidixic acid | 2 (1.5) | - |
| **Nitroimidazole** |  |  |
| Metronidazole | 16 (12.0) | 17 (27.0) |
| **Cephalosporin** |  |  |
| Cefixime | 2 (1.5) | - |
| Ceftriaxone | 1 (0.8) | 1 (1.6) |
| **Thiazolide** |  |  |
| Nitazoxanide | 1 (0.8) | 1 (1.6) |
| **Penicillin** |  |  |
| Pivmecillinum | 1 (0.8) | - |
| Amoxicillin | - | 1 (1.6) |
| **Macrolides + Quinolone** |  |  |
| Azithromycin + Ciprofloxacin | - | 2 (3.2) |
| **Macrolides + Nitroimidazole** |  |  |
| Erythromycin + Metronidazole | - | 1 (1.6) |

The majority of urban children (57.5%) who received antibiotics before coming to the hospital had received them after consultation with a doctor. The scenario is quite the opposite for the rural children, and the difference is statistically significant (p<0.001) (**Table 5**).

**Table 5.** Source of the antibiotics received by 6-23 months old children with rotavirus diarrhea, N= 2801.

| Source | Urban (1923)<br>n (%) | Rural (878)<br>n (%) | P value |
| --- | --- | --- | --- |
| Physician (with prescription) | 1105 (57.5) | 120 (13.7) | p<0.001 |
| Pharmacy (without prescription) | 818 (42.5) | 758 (86.3) |  |

## Discussion

In our study, we observed a significant increase in the proportion of antibiotic use among children with rotavirus diarrhea over the 15-year period from 2004 to 2018, in both urban and rural settings. This finding is consistent with the rising trend of antibiotic consumption reported in many low- and middle-income countries (LMICs), including Bangladesh, as evidenced by large multi-country surveys (28,29).

Several factors might have contributed to this rising trend. One of the key factors responsible might be the less stringent regulation in LMICs allowing over-the-counter sales of antibiotics (30,31). The overall economic growth, as reflected by the increase in the per capita GDP in Bangladesh, could be a further explanation. The increasing GDP of LMIC countries provides for more discretionary spending on high-priced antibiotics (28). Our current study also showed that antibiotic use is significantly higher among those children coming from high-income families (monthly family income, >100US$) in comparison to their low-income counterparts. This difference in the utilization of antibiotics across the different social classes has also been reported by several other studies (32–35). Parental literacy, both paternal and maternal, was found to be independently associated with antibiotic use in children with rotavirus diarrhea in our study. Educated parents often exhibited a greater inclination to treat their children with antibiotics, believing in accelerated recovery. This led to the indiscriminate use of antibiotics, particularly among children of educated parents residing in urban areas (33,34,36,37).

Our study showed that in the urban site, three-fifths of children with rotavirus diarrhea who received antibiotics for their illness visited a physician’s chamber for consultation before coming to icddr,b. Unnecessary practice of antibiotic prescription by physicians has become a concern seen across the LMICs, including Bangladesh (38–40). This physician behavior is likely to be the result of a complex interplay between multiple factors, including inadequate training in the management of viral diarrhea, influence from pharmaceutical companies or employers, peer pressure, and parent expectations of antibiotic treatment, as well as fear of losing patients. (41–45).

Our study found a significantly lower proportion of rural children with viral diarrhea received an antibiotic for their illness. This is probably due to the effect of HDSS maintained by icddr,b for more than 60 years in Matlab; along with the presence of a dedicated diarrheal treatment centre and ongoing counseling through dedicated community health workers. Since its inception, many community-based trials for the treatment of diarrhea (for example, ORS and zinc trials) have taken place in this community (46). Therefore, people in this area might have developed greater awareness about appropriate diarrheal disease management.

The drop in antibiotic use for rotavirus diarrhea in rural Matlab between the period 2008 to 2012 can be explained by two large community-based rotavirus vaccine trials (initiated in April 2007 and continued till June 2011) conducted during the period. These trials covered almost 80% of children aged 0- 23 months living in the Matlab HDSS area. During that time, study field workers visited the homes of children regularly. They encouraged them to bring the child quickly to the Matlab hospital in the event of any diarrheal episode. As a result of this vigilant monitoring, the hospital experienced a surge in the number of children with rotavirus diarrhea during that period (47–49), which might have impacted the decline in antibiotic misuse. The mentioned phenomenon is supported by a systematic review that indicated that mass community vaccination against diseases could reduce the tendency of antibiotic use among the children living in the same community by lowering the incidence of the disease as well as decreasing symptom severity and duration of illness (50).

In our study, living far from a health facility is associated with higher odds of receiving antibiotics for viral diarrhea. This tendency is also reported in other studies conducted in Bangladesh, where this behavior may be explained by poor communication and higher transportation costs, leading caregivers to resort to local practitioners and pharmacies, both of which have a higher propensity to prescribe or dispense antibiotics (21,51). In Bangladesh, retail drug shops or pharmacies are the key healthcare options for 80% of the population (52). Our study also revealed that 87% of the children from the rural area who received antibiotics before coming to the hospital had received them from nearby pharmacies/drug stores. This finding is similar to other reports from this area (53). A study conducted in a similar setting in Bangladesh reported that patients prefer to visit these sources in spite of spending a dime and time behind physician’s prescriptions and laboratory tests. However, unfortunately, half of these drug shops were found to be unauthorized, and most did not have a graduate pharmacist to dispense antibiotics in the first place (52). This ultimately results in incorrect dosing and inappropriate treatment with antibiotics for all diseases treated by those drug sellers. This is a common scenario for many LMICs and has even been reported in a few developed countries (6,54). However, the recently passed law by the Bangladesh government against the over-the-counter sales of antibiotics without prescription might help in bringing down this ongoing tendency of antibiotic overuse (55,56).

Increased frequency (>10 times in the last 24 hours) and longer duration of diarrhea (in hours) were also found to be associated with higher antibiotic use in the current study. Increased frequency and longer duration of rotavirus gastroenteritis, coupled with the absence of any specific medication, sometimes made the parents frightened and frustrated. This feeling of helplessness pushed them to consult physicians and sometimes made them persuade the physicians to treat the illness aggressively with unnecessary antibiotics. Physicians themselves also sometimes get carried away by observing the severity of the symptoms and choose antibiotics for treating rotavirus diarrhea on top of the recommended treatment (21,22,39,57).

In our present study, macrolides appeared to be the main group of antibiotics used in Rotavirus diarrhea, followed by quinolones and nitroimidazole. Macrolides and quinolones are the antibiotics that belong to the “Watch group” of WHO’s Essential Medicines List for Children 2017 (58). In their technical report, WHO recommended using the antibiotics of this group cautiously as they have a higher potential for the development of resistance. In that context, our study findings indicating the abuse of these antibiotics is a matter of great concern for Bangladesh. However, these findings of ours are in line with the recent Lancet publication that also showed that Bangladesh is among the only six countries in the world where the “Watch group” of antibiotics was used in a high proportion (59). Results from the MAL-ED study, as well as a couple of other Bangladeshi studies, including a report by WHO, also support our study findings (20–22).

Rotavirus is the most common cause of diarrhea among children under 2 years of age and is the major contributor to mortality and morbidity in this age group. In order to reduce the burden of rotavirus diarrheal cases, the Government of Bangladesh is planning to introduce the rotavirus vaccine in the National Expanded Program of Immunization (EPI). Nevertheless, previous studies conducted in similar settings have shown that the vaccine can reduce half of the cases (60), which in turn suggests that infection due to rotavirus will prevail up to a certain limit whether we use the vaccine or not. Therefore, we must maintain caution regarding the misuse of antibiotics in Rota viral diarrhea as irrational use of antibiotics in Rota viral infection will intensify the global antimicrobial resistance situation further. The WHO identifies antibiotic resistance as one of the greatest threats to human health in recent times. Ensuring judicious use of antibiotics is a top strategic priority of the WHO’s ‘Global action plan on antibiotic resistance’. Being a member state of the WHO and a key signatory of the ‘Global action plan on antibiotic resistance’, Bangladesh has also recognized the issue of persistent overuse and misuse of antibiotics as an important contributor to the emergence of antibiotic resistance. The country has started working hand in hand with the WHO and other organizations to mitigate the issue (61).

### Strengths and limitations

This study, which to the best of our knowledge is the first study in Bangladesh that systematically investigated 15 years longitudinal trends of irrational antibiotic use in childhood rotaviral diarrhea, shed light on the recent pattern of antibiotic use for the illness and also put an effort to identify the advocating source as well as the factors associated with this behavior both in urban and rural settings. However, it has several limitations. The first was the retrospective nature of the study, which prevented us from systematic data collection and rigorous quality control. Lacking information about the antibiotics belonging to the “Others” group prevented us from reporting their changing trends. Secondly, due to the retrospective nature of our study, we could not follow up with the participants to understand the morbidity caused by the disease and inappropriate antibiotic use. In addition, information regarding caregivers’ and physician’s knowledge and attitude would have been helpful in explaining the study results even further. However, this does not change the objective of this study, which is to investigate the antibiotic misuse, associated factors, and types of antibiotics used among 6-23 months old children presenting with acute rotavirus diarrhea.

## Conclusion and recommendations

The present study has identified that antibiotic use in children with rotavirus diarrhea has been increasing steadily over the years and that many physicians were involved in this practice of prescribing antibiotics irrationally. Besides, the present study also reported that Macrolides, followed by Quinolones and Nitroimidazole, were the main groups of antibiotics being used recently in the illness. These findings of our study are, in fact, very alarming for a country like Bangladesh, which is already under the threat of emerging antimicrobial resistance. In addition, the current study has identified several sociodemographic as well as epidemiologic factors, including higher family income, maternal & paternal literacy, residence distant from nearby health facilities, and increased frequency and duration of diarrhea to be associated with this growing practice. All these findings of our study warrant the necessity of design campaigns targeting both the patients/caregivers, physicians, and other relevant health personnel to raise their awareness about the grave consequences of injudicious use of antibiotics. Furthermore, the results of our study also demand continuous vigilant surveillance and stringent implementation of the existing law to reduce the indiscreet use of antibiotics in Bangladesh.

## Data Availability

All aggregated data related to this study are provided in the paper. To protect the identification of patients derived from the composite of key study variables, some restrictions do apply to the primary data. These data can be made available from the Ethics Committees (RRC and ERC) of icddr,b for researchers who meet the criteria for access to confidential data. Readers may contact the Senior Manager of Research Administration at icddr,b (Shiblee Sayeed) for data policies and queries.

## Acknowledgment

Hospital surveillance activity was funded by core donors who provide unrestricted support to icddr,b for its operations and research. Current donors providing unrestricted support include the Governments of Bangladesh and Canada. We gratefully acknowledge our core donors for their support and commitment to icddr,b’s research efforts. We also acknowledge the support of Dr. Amena Al Nishan and Dr. Syed Imran Ahmed from icddr,b and Dr. Charles P Larson from McGill University Global Health Programs in conducting the study.

## Funding

No funding was allocated for this specific work.

## Conflict of Interest

The authors have declared that no competing interests exist with respect to the research, authorship, and publication of this article.

## Author Contributions

Conceptualization: Kazi Nazmus Saqeeb, S. M. Tafsir Hasan

Data curation: Soroar Hossain Khan

Formal analysis: Soroar Hossain Khan, Kazi Nazmus Saqeeb

Investigation: Soroar Hossain Khan, ASG Faruque

Methodology: S. M. Tafsir Hasan, Kazi Nazmus Saqeeb

Project administration: ASG Faruque, Tahmeed Ahmed, Md. Alfazal Khan

Resources: Tahmeed Ahmed

Software: Soroar Hossain Khan

Supervision: ASG Faruque, Tahmeed Ahmed

Validation: S. M. Tafsir Hasan, ASG Faruque

Visualization: Kazi Nazmus Saqeeb, Soroar Hossain Khan

Writing – original draft: Kazi Nazmus Saqeeb, S. M. Tafsir Hasan

Writing – review & editing: ASG Faruque, Tahmeed Ahmed, Md. Alfazal Khan

## Notes

### Competing Interest Statement

The authors have declared no competing interest.

### Funding Statement

The author(s) received no specific funding for this work.

### Author Declarations

The Diarrheal Disease Surveillance System (DDSS) is an integral component of the ongoing operations at icddr,b's Dhaka and Matlab hospitals. Informed verbal consent was obtained from all prospective patients or their legal guardians/caregivers at the time of enrollment. This consent was documented by checking the appropriate box on the questionnaire and was reviewed with the participants or their parents for confirmation. Participants were properly informed of the voluntary nature of their participation and their right to withdraw at any time. The confidentiality of the information collected was assured, and participants were informed that future analyses and publications would be conducted anonymously. The collected information was explained to be used for improving patient care. This surveillance process has been approved by the Research Review Committee (RRC) and Ethics Review Committee (ERC) of icddr,b.

